# A Cell-Based Assay for Detection of Anti-Fibrillarin Autoantibodies in Systemic Sclerosis

**DOI:** 10.1101/2022.06.07.22276113

**Authors:** Gerson Dierley Keppeke, Minoru Satoh, Cristiane Kayser, Pedro Matos, Tomoko Hasegawa, Shin Tanaka, Larissa Diogenes Santos, Rogerio Quintiliano Amaral, Silvia Helena Rodrigues, Luis Eduardo Coelho Andrade

## Abstract

**Objectives:** Anti-fibrillarin antibodies are useful for establishing diagnosis and predicting distinct clinical features in systemic sclerosis (SSc). Anti-fibrillarin produces a characteristic clumpy nucleolar pattern in indirect immunofluorescence assay on HEp-2 cells (HEp-2 IFA) that is useful to guide further testing in antigen-specific immunoassays (ASI). Immunoprecipitation (IP) is the gold standard ASI for anti-fibrillarin determination. We established a new anti-fibrillarin Cell-Based Assay (CBA) and compared its diagnostic performance with IP and ASI commercial kits. The clinical features of SSc patients with and without anti-fibrillarin antibodies were analyzed.

**Methods:** A TransMembrane Signal (TMS) was added to the human fibrillarin gene in order to drive the transgenic fibrillarin to the cytoplasmic membrane. HEp-2 cells transfected with a vector containing the *TMS-fibrillarin* were used as substrate for IFA in the CBA. Sixty-two samples with high-titer nucleolar pattern in HEp-2 IFA (41 clumpy and 21 homogeneous/punctate) were tested for anti-fibrillarin in the CBA, IP, line-blot and ELISA. Clinical SSc phenotype was evaluated in 106 patients according to positive/negative anti-fibrillarin result in the CBA.

**Results:** TMS-fibrillarin was properly located to the cytoplasmic membrane and recognized by human autoantibodies. Thirty-eight of 41 clumpy nucleolar samples (92.7%) and none of 21 samples with other nucleolar patterns were positive for anti-fibrillarin in the CBA. There was 100% agreement between the positive/negative results in the CBA and IP. Among the 38 CBA-positive samples, only 15 (39.5%) and 11 (29%) were considered positive for anti-fibrillarin in the line-blot and ELISA, respectively. Anti-fibrillarin was associated with higher frequency of diffuse cutaneous SSc (dcSSc) phenotype (72.7% vs 36.8%; p=0.022), cardiac involvement (36.4% vs 6.5%; p=0.001) and scleroderma renal crisis (18.2% vs 3.3% p = 0.028).

**Conclusion:** With an innovative strategy of targeting the transgenic autoantigen to the cell membrane, we developed a new straightforward assay for detection of anti-fibrillarin autoantibodies. This new CBA presented high sensitivity and specificity for the detection of anti-fibrillarin autoantibody, comparable to the gold standard IP. Moreover, anti-fibrillarin antibodies detected in the CBA identified patients with a higher frequency of dcSSc, cardiac and renal involvement.

## Introduction

Systemic sclerosis (SSc) is a chronic heterogeneous systemic autoimmune disease characterized by microvascular dysfunction, activation of the immune system, and cutaneous and visceral fibrosis. SSc has the highest morbidity and mortality rates among immune-mediated rheumatic diseases [1]. However, the clinical manifestations and the clinical course of the disease are highly variable. The heterogeneity of the disease may be represented by the subset classification of SSc, i.e., the limited (lcSSc) and diffuse cutaneous subsets (dcSSc). Patients with dcSSc have rapidly diffuse cutaneous thickening, higher frequency of internal organ involvement, and worse prognosis. In contrast, patients with lcSSc typically have restricted skin thickening distribution, less severe organ involvement and a better survival [2].

Serum autoantibodies can be detected in over 90% of SSc patients and are very useful for the early diagnosis of SSc and for the identification of certain SSc disease phenotypes. Several of these autoantibodies are highly specific for SSc and help predicting clinical complications and the prognosis of SSc patients [3, 4]. For example, anti-topoisomerase I antibodies are associated with dcSSc, digital ulcers, cardiomyopathy, high skin score, higher risk of severe interstitial lung disease (ILD) [5] and underlying malignancies [6], resulting a more severe phenotype and increased mortality [4, 7-9]. Anti-RNA polymerase III (RNAP III) antibodies are associated with the dcSSc subset, high risk of severe, rapidly progressing cutaneous thickening, higher risk for gastric antral vascular ectasia [10, 11], and scleroderma renal crisis [10, 12], as well as a higher risk of development of malignancy, especially within the first years of disease onset [6, 10]. In contrast, anti-centromere antibodies are associated with lcSSc, long-standing Raynaud’s phenomenon, calcinosis, a higher risk of developing pulmonary arterial hypertension (PAH) [13], and better survival rates as compared with patients with anti-topo I and anti-RNAP III antibodies [7].

Anti-U3-RNP/fibrillarin antibodies recognize the U3-ribonucleoprotein (U3-RNP), a nucleolar complex involved in pre-rRNA processing. Anti-fibrillarin occurs in up to 10% of SSc patients, especially in African-Americans [14, 15]. Although there are not many studies, anti-fibrillarin has been associated with dcSSc and multi-organ involvement [9, 16], with high risk of cardiac involvement, ILD, PAH, renal crisis, small bowel and muscle involvement. In general, anti-fibrillarin antibodies indicate worse prognosis in SSc patients [9, 14].

The indirect immunofluorescence assay on HEp-2 cells (HEp-2 IFA) is the most commonly used method for screening for autoantibodies in systemic autoimmune diseases, including SSc. One key information of the HEp-2 IFA test is the immunofluorescence pattern, which indicates the possible location of the autoantigens recognized by the autoantibodies in the sample. The International Consensus on ANA Patterns (ICAP) <www.anapatterns.org> has classified 30 different HEp-2 IFA patterns [17]. At the expert level, the nucleolar pattern can be distinguished into three different “subpatterns”: homogeneous nucleolar AC-8, clumpy nucleolar AC-9, and punctate nucleolar AC-10, respectively. The nucleolar pattern, usually at high titer, is observed in up to a third of SSc patients [18], although in most studies the proportion of this pattern is around 20% [19-22]. A comprehensive review on the topic can be found elsewhere [23]. A portion of samples from patients with SSc with a nucleolar HEp-2 IFA pattern have anti-fibrillarin (U3-RNP) antibodies, which typically yield a clumpy nucleolar pattern (AC-9) [4]. However, the correct classification of the clumpy nucleolar pattern can be challenging as the interpretation is subjective, depends on qualified personnel, and may vary depending on the brand of the HEp-2 slides [24]. In addition, is possible that not all clumpy nucleolar patterns are caused by anti-fibrillarin antibodies.

Therefore, when a sample presents a nucleolar pattern in the HEp-2 IFA test, additional tests should be perform to confirm the presence of anti-fibrillarin or other autoantibodies that could yield the pattern [4]. There are commercial kits for anti-fibrillarin antibodies based on solid phase immunoassays, such as western-blot, dot-blot, line-blot, fluorescence enzyme immunoassay, ELISA, particle-based multi-analyte technology, among others [25, 26]. However, most of these kits are for Research Use Only (RUO). In addition, they can yield inconsistent results with poor sensitivity, probably because of difficulties in the preservation of the native conformation of fibrillarin. Therefore, immunoprecipitation (IP) is considered the gold standard method for detection of anti-fibrillarin antibodies [23]. Unfortunately, IP is a labor-intensive assay that requires highly skilled analysts and the use of radioactive materials; thus, this method is not widely available in routine clinical laboratories.

In this study, we established an indirect immunofluorescence Cell-Based Assay (CBA) to detect anti-fibrillarin antibodies and compared the performance of this new assay against standard IP and commercial solid phase immunoassays for detection of anti-fibrillarin autoantibody. In addition, we surveyed a cohort of SSc patients with the novel anti-fibrillarin CBA test and confirmed previously observed phenotypic associations of anti-fibrillarin antibodies.

## Results

A TransMembrane Signal (TMS) was fused to the N’ terminus to the human fibrillarin gene so that the transgenic fibrillarin is guided to the cytoplasmic membrane. HEp-2 cells transfected with the plasmid *pCMV_TMS-fibrillarin_P2A_OFP-myc* were used as substrate for IFA in the CBA. Indirect Immunofluorescence (IIF) labeling with an anti-human fibrillarin mouse monoclonal antibody indicates that the TMS-Fibrillarin was successfully positioned at the cell membrane (Suppl. Figure 1A). Cells expressing TMS-fibrillarin could be readily identified by the Orange Fluorescent Protein (OFP) in the construct (TMS-fibrillarin_P2A_**OFP**-myc). However, since fixing with methanol/acetone affects OFP fluorescence, labeling of the transfected cells was enhanced using an anti-myc tag monoclonal antibody (Suppl. Figure 1B). As expected, the cell monolayer contains transfected and non-transfected cells side by side.

In the IIF cell-based assay (Fibrillarin/CBA), the cytoplasmic membrane of the cells expressing TMS-Fibrillarin was stained by two human sera that present a clumpy nucleolar pattern (AC-9) in the HEp-2 IFA test, but not by a serum that presents homogeneous nucleolar (AC-8) pattern in the HEp-2 IFA test (Figure 1). No cytoplasmic membrane staining in the Fibrillarin/CBA was observed using samples that yield a speckled nucleolar pattern (AC-10) in the traditional HEp-2 IFA test (data not shown). To validate the assay, we selected 62 samples with strong nucleolar pattern on the HEp-2 IFA test from the routine in the clinical laboratory. Antibody titer and the nucleolar pattern (homogeneous nucleolar AC-8, clumpy nucleolar AC-9, or punctate nucleolar AC-10) were confirmed with traditional HEp-2 IFA slides (Euroimmun). Classification of the nucleolar patterns followed the recommendations from the International Consensus on ANA Patterns (ICAP) and samples with titer≥1/320 were included. Representative images of samples with the different nucleolar patterns are shown in Suppl. Figure 2A-E.

**Figure 1.**
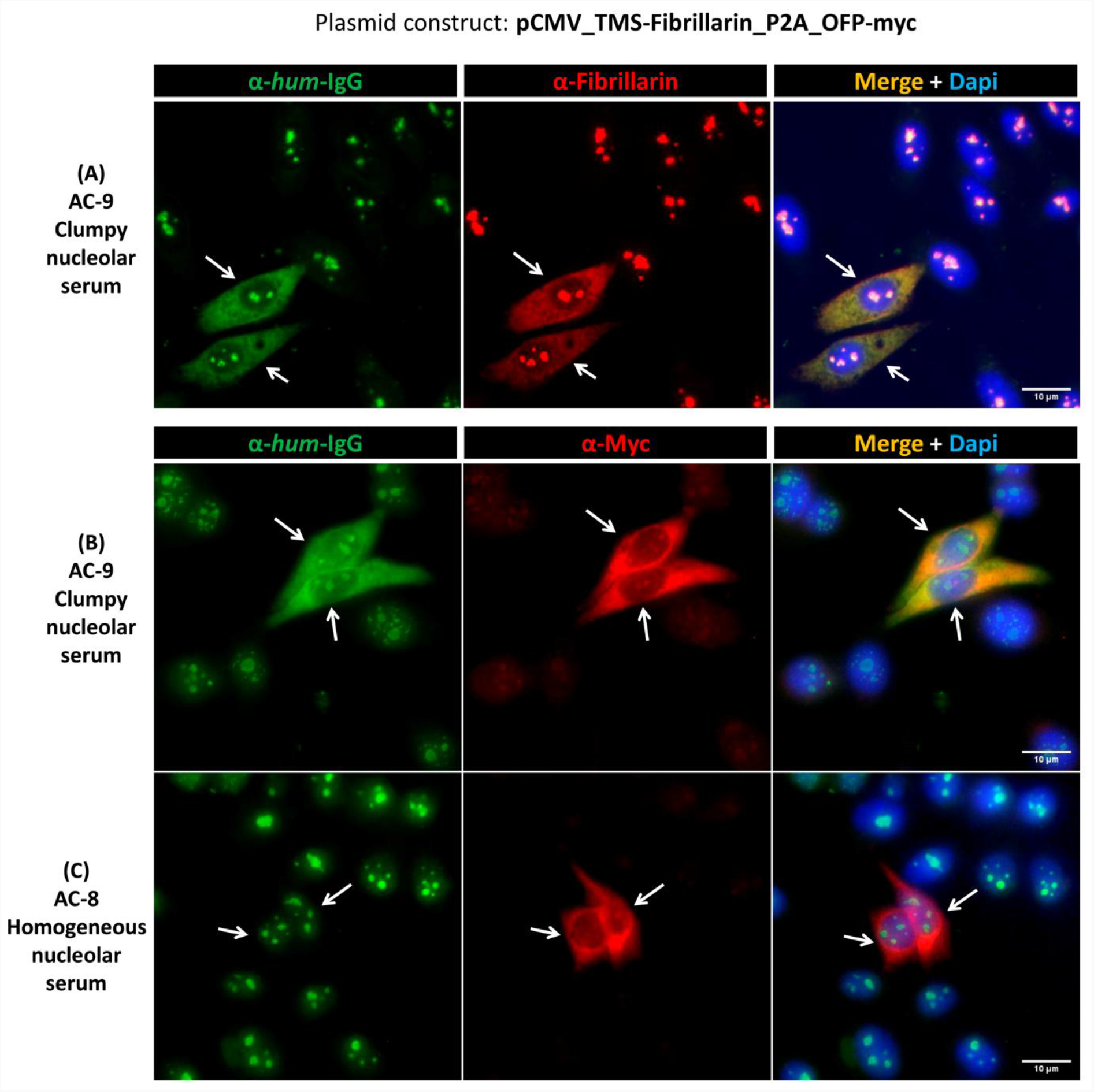
Labeling of TMS-Fibrillarin by serum with clumpy nucleolar pattern in HEp-2 IFA. HEp-2 cells transfected with the TMS-Fibrillarin plasmid were labeled with human serum plus fluorescent mouse monoclonal antibodies. (A-B) Double IIF using human serum with clumpy nucleolar pattern (AC-9) plus mouse anti-fibrillarin (A) or anti-myc (B) monoclonal antibodies indicates that human antibodies recognize TMS-Fibrillarin at the cell membrane. (C) Serum with homogenous nucleolar pattern does not label the cytoplasmic membrane in cells transfected with TMS-Fibrillarin. Arrows indicate TMS-Fibrillarin expressing cells. Scale bar = 10µm.

We intentionally enriched the collection of samples with the clumpy nucleolar (AC-9) pattern. Therefore, from the 62 samples with the nucleolar pattern, 41 (66%) were classified as clumpy nucleolar (AC-9) and 21 (34%) as other nucleolar patterns (20 as AC-8 and 1 as AC-10). Thirty-eight of 41 AC-9/clumpy nucleolar samples (92.7%) and none of 21 samples with other nucleolar patterns were positive in the Fibrillarin/CBA (Table 1). Representative images of samples that were positive and negative, respectively, in the Fibrillarin/CBA test are shown in Figure 1 and Suppl. Figure 3. Samples that were positive in the Fibrillarin/CBA test showed higher HEp-2 IFA titer than samples that were negative (Suppl. Figure 2F).

**Table 1.** Reactivity in the Fibrillarin /CBA test and other methods for determination of anti-fibrillarin antibodies in 62 serum samples with high-titer reactivity to the nucleolus in HEp-2 IFA.

| Samples (n=62) |  | Fibrillarin/CBA test |  | P |
| --- | --- | --- | --- | --- |
|  |  | Positive<br>38 (61.3%) | Negative<br>24 (38.7%) |  |
| HEp-2 IFA nucleolar pattern | Clumpy nucleolar (AC-9) (n=41; 66%) | 38 (92.7%) | 3 (7.3%) | - |
|  | Homogeneous or punctuate nucleolar (AC-8 and AC-10) (n=21; 34%) | 0 (0%) | 21 (100%) |  |
| Immunoprecipitation fibrillarin reactivity (n=56) | Positive (n=35; 62.5%) | 35 (100%) | 0 (0%) | p<0.001 |
|  | Negative (n=21; 37.5%) | 0 (0%) | 21 (100%) |  |
| Line blot for anti-fibrillarin (Euroline) (n=62) | Positive (++, +++) (n=16; 25.8%) | 15 (93.7%) | 1 (6.3%) | p=0.002 |
|  | Neg / *borderline* (0, +) (n=46; 74.2%) | 23 (50%) | 23 (50%) |  |
| ELISA for anti-fibrillarin (n=62) | Positive (O.D. $\geq 2.1$) (n=12; 19.4%) | 11 (91.7%) | 1 (8.3%) | p=0.016 |
| | Negative (O.D. $\leq 2.0$) (n=50; 80.6%) | 27 (54%) | 23 (46%) | |

Next, we compared the results obtained in the Fibrillarin-CBA test and in the immunoprecipitation (IP) for 56 of the 62 samples with nucleolar reactivity (six samples had not enough remaining volume) (Suppl. Figure 4). Out of the 56 samples, 35 (62.5%) were positive and 21 (37.5%) were negative for anti-fibrillarin antibodies, with 100% agreement of the positive/negative results between the Fibrillarin/CBA test and IP (Table 1). We also analyzed the presence of anti-fibrillarin using the Euroline Systemic Sclerosis Line Blot (Euroimmun) (Suppl. Figure 5). Out of the 62 samples, 16 (25.8%) were positive and 46 (74.2%) were negative for anti-fibrillarin (Table 1). From the 38 samples that were positive in the Fibrillarin/CBA assay, only 15 (39.5%) were considered positive in the line blot assay (Figure 2A). From the 24 samples negative in the Fibrillarin/CBA, one was positive in the line blot assay (Figure 2A). Finally, we analyzed the presence of anti-fibrillarin with a commercial ELISA test for anti-fibrillarin antibody (MyBioSource). Out of the 62 samples, 12 (19.4%) were considered positive according to the manufacturer’s recommendation for defining the cutoff (Table 1 and Figure 2B). Fifty samples (80.6%) were considered negative. Among the 38 samples with positive result in the Fibrillarin/CBA test, only 11 (29%) were positive in the ELISA test (Figure 2B). Among the 24 samples with negative result in the Fibrillarin/CBA, one was positive in the ELISA (Figure 2B).

**Figure 2.**
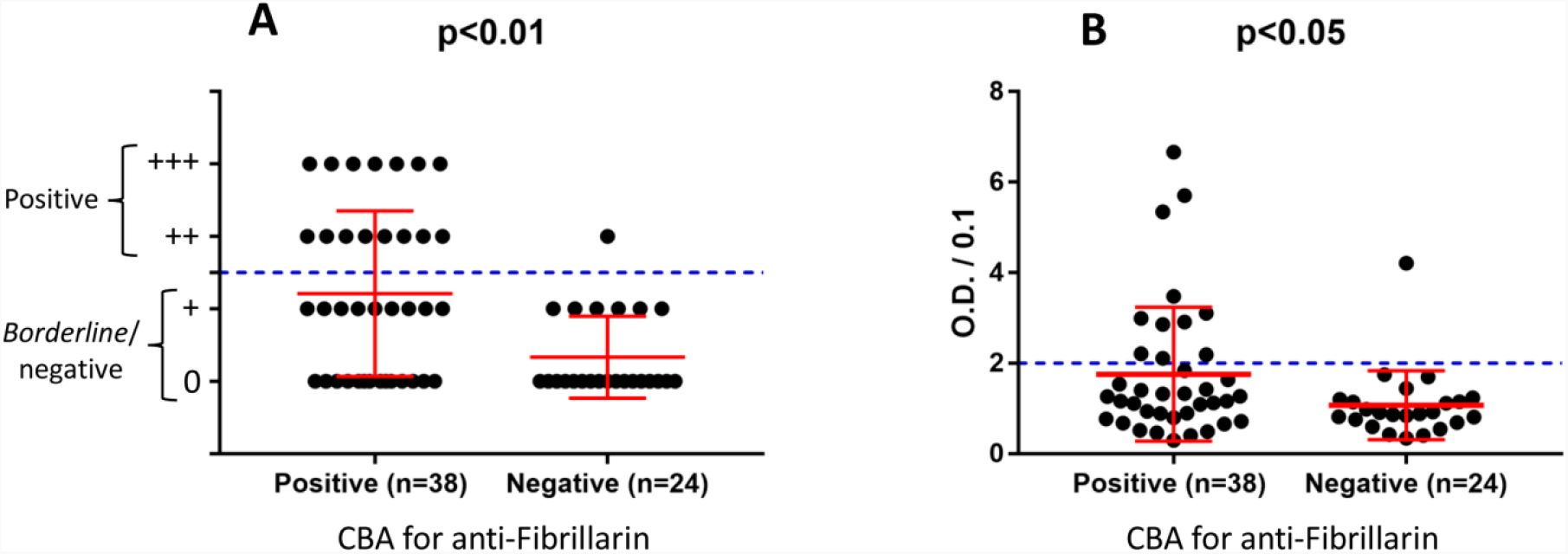
Performance of the Fibrillarin/CBA test and other methods for determination of anti-fibrillarin antibodies. (A) Line blot assay for anti-fibrillarin antibody. The plus (+) symbols in the y-axis indicate the intensity of staining of the fibrillarin line; ≥2 plus (++) was considered positive. (B) ELISA test for anti-fibrillarin antibody. Optic Density (O.D.) was divided by 0.1 and values >2 were considered positive as recommended by the manufacturer. Error bars indicate mean ± SD. Statistics by Mann Whitney test.

For the clinical validation of the Fibrillarin/CBA test, we evaluated a cohort with 106 patients classified as SSc according to the ACR/EULAR SSc classification criteria [27]. The HEp-2 IFA pattern was defined according the ICAP recommendations using commercial HEp-2 slides (Table 2). Thirty-three patients (31.1%) presented nucleolar pattern (AC-8, AC-9, AC-10). Altogether, the most common pattern was the nuclear fine speckled AC-4 pattern (38.7%), followed by nucleolar (31.1%) and by centromere AC-3 (13.2%). Some samples presented more than one pattern. Nine samples (8.5%) were negative in the HEp-2 IFA test. Eleven samples (10.4% from the total) were positive in the Fibrillarin/CBA test. All these 11 samples belonged to the group of 33 samples with nucleolar pattern in the HEp-2 IFA, meaning one third (33.3%) of the samples with nucleolar pattern were positive in the Fibrillarin/CBA assay. None of the 73 samples with other patterns or negative in the HEp-2 IFA test showed reactivity in the Fibrillarin/CBA test (Table 2), suggesting that to be considered positive for anti-fibrillarin a sample must first present nucleolar pattern in the HEp-2 IFA test.

**Table 2.** Anti-fibrillarin reactivity and HEp-2 IFA patterns in 106 systemic sclerosis (SSc) patients.

| HEp-2 IFA pattern | Number (%)<br>Total of 106 <sup>#</sup> | Positive in the<br>Fibrillarin/CBA test |
| --- | --- | --- |
| Nucleolar (all) (AC-8, AC-9, AC-10) | 33 (31.1%) | 11 (10.4%) |
| Centromere (AC-3) | 14 (13.2%) | 0 |
| Nuclear fine speckled (AC-4) | 41 (38.7%) | 0 |
| Nuclear coarse speckled (AC-5) | 6 (5.7%) | 0 |
| DNA topoisomerase I-like (AC-29) | 9 (8.5%) | 0 |
| Other patterns* | 13 (12.3%) | 0 |
| Negative (AC-0) | 9 (8.5%) | 0 |

\*Nuclear homogeneous (AC-1), NuMA-like (AC-26), Cytoplasmic dense fine speckled (AC-19), Cytoplasmic discrete dots (AC-18), nuclear envelope (AC-11/AC-12).
<sup>#</sup> Some samples presented overlapping patterns.

Demographic and clinical features of the 106 SSc patients are described in Table 3. SSc patients had a mean age of 50.2 ± 13.2 years, were mainly females (88.7%), with mean disease duration of 6.7 ± 5.7 years. Patients with positive Fibrillarin/CBA test had mostly dcSSc (72.7%), compared to patients with negative Fibrillarin/CBA test (36.8%) (p = 0.022). A positive Fibrillarin/CBA test was associated with a higher frequency of cardiac involvement and scleroderma renal crisis compared to those with negative Fibrillarin/CBA test (36.4% versus 6.5%, p=0.001; 18.2% versus 3.3%, p=0.028, respectively).

**Table 3.** Demographic and clinical data of systemic sclerosis (SSc) patients according to reactivit in the Fibrillarin/CBA test.

| Variable | Fibrillarin/CBA<br>positive<br>(n = 11) | Fibrillarin/CBA<br>negative<br>(n = 95) | *P* |
| --- | --- | --- | --- |
| Age, mean ± SD (years) | 45.2 ± 14.1 | 50.8 ± 13.1 | 0.182 |
| Female/Male, n (%) | 10 (90.9) /1 (9.1) | 84 (88.4) /11 (11.6) | 0.805 |
| Disease subset, n (%) |  |  |  |
| lcSSc | 3 (27.3) | 60 (63.2) | 0.022 |
| dcSSc | 8 (72.7) | 35 (36.8) |  |
| Disease duration, mean ± SD (years) | 5.4 ± 3.8 | 6.8 ± 5.9 | 0.652 |
| **Organ involvement** |  |  |  |
| Digital ulcers, n (%) | 5 (45.5) | 44 (47.3) (n=93) | 0.907 |
| Esophageal dysmotility, n (%) | 8 (72.7) | 80 (86.0) (n=93) | 0.248 |
| Small bowel involvement | 2 (18.2) | 9 (9.7) (n=93) | 0.386 |
| FVC % of predict, mean ± SD | 72.2 ± 24.8 | 79.4 ± 18.9 | 0.306 |
| ILD, n (%) | 6 (54.5) | 56 (59.6) (n=94) | 0.748 |
| PAH, n (%) | 1 (9.1) | 14 (15.0) (n=93) | 0.595 |
| Cardiac involvement, n (%) | 4 (36.4) | 6 (6.5) (n=93) | 0.001 |
| Scleroderma renal crisis, n (%) | 2 (18.2) | 3 (3.3) (n=93) | 0.028 |
| Arthritis or myositis, n (%) | 4 (36.4) | 29 (31.2) (n=93) | 0.727 |

dcSSc: diffuse cutaneous SSc; lcSSc: limited cutaneous SSc;
FVC: forced vital capacity; ILD: interstitial lung disease; PAH: pulmonary arterial hypertension.

## Discussion

SSc is a severe systemic autoimmune disease and the identification of SSc-specific autoantibodies is important not only for diagnosis but also for the prediction of clinical manifestations and outcome. In particular, the identification of anti-fibrillarin antibody is helpful because it is associated with a more severe disease and higher morbidity and mortality. In the present study we originally developed a new CBA for the detection of anti-fibrilarin autoantibodies that showed sensitivity and specificity compared to the gold standard IP test.

Solid-Phase Immunoassays (SPIA) for cell-membrane protein antigens usually present poor performance regarding sensibility and specificity, most likely because the antigen loses its natural conformation when removed from the lipid membrane microenvironment. Thus, for some assays we resort to animal tissue sections as substrate [28] and for others the best current option is still immunoprecipitation with radiolabeled cell extracts. However, the latter method is not practical for routine clinical laboratories worldwide. Recent developments have demonstrated CBA technology to be an efficient strategy to replace or complement IFA on animal tissue sections and immunoprecipitation for detection of disease-relevant specific autoantibodies in patient samples. Application of CBA for clinical immunodiagnostics have increased in recent years, with prospect for further increase. As an example, the recent expansion in the field of neuroimmunology has largely benefited from this methodological platform as specific autoantibody biomarkers area readily determined by IFA on cells transfected with aquaporin-4, N-methyl-D-aspartate receptor (NMDAR), Muscle-Specific Kinase (MuSK), Myelin oligodendrocyte glycoprotein (MOG), among others [28]. Likewise, CBA have been successfully used to determine autoantibodies against Phospholipase A2 Receptor (PLA2R) and Thrombospondin type 1 domain–containing 7A (THSD7A), both biomarkers of primary membranous glomerulopathy [29, 30].

Typically, the CBA methodology uses eukaryotic cells transfected with a vector (usually plasmids) carrying the gene for the protein of interest. The chosen cell line should not naturally express the given autoantigen, allowing for the use of non-transfected cells as negative control. The transfection will “force” the cells to express the antigen, allowing them to be used as substrate in an indirect immunofluorescence reaction where the autoantibodies present in the patient serum are the primary probe.

In this study, we employed an innovative strategy, where the gene was engineered so the product of the fibrillarin transgene could be relocated at a different cellular site from the constitutive fibrillarin. The TransMembrane Signal fused to the N-terminus of the fibrillarin transgene causes the protein to be ultimately expressed in the extracellular surface of the cytoplasmic membrane, in a “receptor-like” fashion. The transfected and non-transfected cells were used as substrate for a standard IFA, which displayed sensibility and specificity for detection of anti-fibrillarin antibodies equivalent to that of the gold standard IP assay. It is likely that such a good performance occurred because the endogenously synthesized transgenic fibrillarin was presented at the cell surface in its native-like conformation. This result encourages future studies applying a similar strategy to develop CBAs for detection of autoantibodies to other autoantigens, especially those where traditional SPIA display suboptimal analytical and diagnostic performance.

Previous studies have shown that the nucleolar pattern may account for ∼5% of all positive HEp-2 IFA results [31-34]. The nucleolar pattern, especially in high titer, is also classically associated with SSc, and the proportion of nucleolar positive samples in SSc varies from 10% up to 40%, depending on the ethnicity and disease subsets evaluated [22, 23]. In our study, we found that 31.1% of the samples presented nucleolar pattern in the HEp-2 IFA test (some samples had additional overlapping patterns), and one third of the samples with nucleolar pattern (33.3%) where positive for anti-fibrillarin, representing 10.4% of the total SSc cohort. This frequency is similar to previous studies that have shown a prevalence of anti-fibrillarin reactivity ranging from 5 to 10% of SSc [14, 15, 23, 26].

In agreement with previous studies, we found a higher frequency of diffuse cutaneous involvement [9] and higher frequency of cardiac and renal involvement [35, 36] among patients with anti-fibrillarin autoantibodies, indicating a more severe disease among these patients. However, we did not find association with GI involvement [35], ILD and PAH [14], as observed in other cohorts of SSc patients [4]. The relatively small number of patients in the present cohort may have prevented the identification of association with these manifestations. In a study of the Pittsburgh Scleroderma Databank, evaluating 1,432 patients with SSc, anti-fibrillarin was associated with dcSSc and multi-organ involvement including joint involvement, severe gastrointestinal disease, pulmonary fibrosis, PAH, digital ulcers and heart and kidney involvement [9]. Of note, a high proportion of these patients were African-Americans, what could explain some of the different results observed in our patients, who had different ethnic background. Moreover, another study evaluating African-American patients with SSc also found a higher frequency of digital ulcers, GI involvement, and pericarditis but less severe pulmonary involvement among patients that were anti-fibrillarin positive [15]. Apart of some phenotype differences among the several studies, all indicate a more severe disease and decreased survival rate among patients with anti-fibrillarin antibodies [37].

Our data indicate that anti-fibrillarin antibody results based on traditional SPIA in the clinical laboratory must be taken with caution. Among the 38 Fibrillarin/CBA-positive samples, only 15 (39.5%) and 11 (29%) were considered positive in the line-blot and ELISA, respectively. The poor sensitivity of the line-blot was a surprise, especially since a previous study found that line immunoblot results for detection of anti-fibrillarin has comparable clinical significance with those of immunoprecipitation results [26]. SPIA specificity was reasonable in our study. Among the 24 samples with negative result in the Fibrillarin/CBA, one was positive in the line blot assay and one was positive in the ELISA. These results could be considered a false-positive. Of note, the false-positive sample in the line-blot was not the same as the false-positive by the ELISA.

One limitation of the herein described Fibrillarin/CBA is the possible interference of other autoantibodies in the sample. In our model, TMS-Fibrillarin is relocated to the cell membrane and the read-out of the IFA in positive samples is an immunofluorescence pattern in the transfected cells that resembles a dense fine speckled cytoplasmic pattern (Figure 1 and Suppl. Figure 3). If a given sample presents autoantibodies against a cytoplasmic autoantigen, the interpretation of the Fibrillarin/CBA may be affected. In fact, from the 106 samples of SSc patients, seven (6.6%) had an “inconclusive” result at the initial 1/80 screening dilution in the Fibrillarin/CBA IFA reaction, including four samples with dense fine speckled cytoplasmic pattern (AC-19) in standard HEp-2 IFA. None of those seven presented nucleolar pattern in the standard HEp-2 IFA, and they were all classified as negative for anti-fibrillarin by the CBA after further testing with serial 2x dilution. In such cases, it is possible that sequential double dilution can be helpful in samples in which the anti-fibrillarin titer is higher than the titer of the concurrent anti-cytoplasm antibodies. Along the sequential dilution process, the TMS-fibrillarin staining of transfected cells would have higher intensity than the progressively fainter staining of non-transfected cells. On the other hand, such double dilution process cannot exclude anti-fibrillarin reactivity in samples with equivalent reactivity intensity of concomitant anti-cytoplasmic antigen.

## Conclusion

In this proof-of-concept study, we present an innovative strategy, i.e., the relocation of the transgenic autoantigen to a cell topographic site different from that of the endogenous autoantigen. This strategy overcomes one limitation in the development of CBA for autoantibodies against autoantigens regularly expressed in most cell types. The herein presented Fibrillarin/CBA is cost-effective and as easy to perform as a standard HEp-2 IFA reaction, rendering it ready for adoption at clinical immunodiagnostic laboratories. The performance of the Fibrillarin/CBA method was comparable to the gold standard immunoprecipitation method and had higher sensitivity than commercially available anti-fibrillarin antibody solid-phase assay technologies. In addition to the successful analytical validation, the Fibrillarin/CBA depicted the expected clinical performance, being associated with severe manifestations of SSc including diffuse cutaneous involvement, cardiac and renal involvement.

## Materials and methods

### Patient samples

Sixty-two samples with nucleolar pattern and titer ≥1/320 in the HEp-2 IFA test were used to validate the fibrillarin CBA. These samples were sequentially selected from the routine HEp-2 IFA operation at the laboratory of the Rheumatology Division, Department of Medicine, Federal University of Sao Paulo and at the Fleury clinical laboratory. No identification data or clinical information was available for these samples. Considering that the samples were used exclusively for immunoassays related to the original physician’s prescription and no identification or clinical information was used, the Ethics Committee waived the need for informed consent for these 62 samples. An additional set of samples from a cohort of 106 patients meeting the American College of Rheumatology/European League Against Rheumatism (ACR/EULAR) 2013 classification criteria for SSc [27] was tested with the anti-fibrillarin CBA test, aiming to determine the frequency and clinical associations of anti-fibrillarin antibodies. The patients were consecutively recruited from the Rheumatology outpatient clinic at the Federal University of Sao Paulo and had their electronic medical records thoroughly reviewed by rheumatologists with expertise in SSc (C.K. and P.M.). The patients signed an informed consent form to participate in the study (the research was approved by Local Ethics Committee at the Federal University of Sao Paulo).

Clinical and demographic characteristics of SSc patients including age, gender, disease subtype (limited or diffuse cutaneous SSc), and disease duration (defined as the time between the first non-Raynaud’s symptom and the last available evaluation) were obtained from medical records as previously described [38]. Briefly, the presence of esophageal dysmotility was evaluated by barium esophagogram or esophageal manometry and/or endoscopy. The presence of small bowel involvement was considered in cases with small intestinal bacterial overgrowth characteristic symptoms, including bloating, abdominal distension, diarrhea, and weight loss. Pulmonary evaluation included pulmonary function test (PFT), chest high resolution computed tomography (HRCT) and echocardiography performed within the last 12 months. Interstitial lung disease (ILD) was defined by the presence of any radiologic evidence of interstitial abnormalities on HRCT and forced vital capacity (FVC) <80% in PFT. Pulmonary arterial hypertension (PAH) was confirmed by right heart catheterization and defined as a mean pulmonary arterial pressure (mPAP) ≥25 mmHg with a pulmonary artery wedge pressure ≤15 mmHg and pulmonary vascular resistance >3 Wood units [39, 40]. Cardiac involvement included arrhythmias, myocardial dysfunction and/or myocardiosclerosis documented by an electrocardiogram, echocardiogram or cardiac magnetic resonance imaging. History or presence of active digital ulcers (DU), scleroderma renal crisis (SRC) and musculoskeletal involvement (arthritis or myositis) was also registered.

### Plasmid cloning

Total RNA was extracted from human peripheral blood mononuclear cells using TRIzol (Invitrogen, EUA). The RNA was converted to complementary DNA (cDNA) with the Kit “First Strand cDNA Synthesis Kit” (E6300, NEB, EUA). From the cDNA, the coding region of fibrillarin gene *FBL* (NCBI Reference Sequence: NM_001436.4) was amplified with the following primers (Forward: ATGAAGCCAGGATTCAGTCCC; Reverse: GTTCTTCACCTTGGGGGGTG), using the Phusion Flash High-Fidelity PCR Master Mix (F548, Thermo Scientific, EUA). Size and sequence were confirmed by agarose gel and Sanger sequencing, respectively.

The TransMembrane Signal (TMS) was added to the N-terminus of the fibrillarin gene. TMS is a proprietary 65-amino acids sequence designed by the study authors (GDK and LECA) aiming to guide the gene product to the cell membrane. The cDNA for the TMS fragment was synthesized by TsingKe Biological Technology (China). At the C-terminus of the fibrillarin gene, an orange fluorescent protein (OFP) gene plus a fused myc tag was added, interleaved from *FBL* by a ribosome skipping P2A sequence. The whole cDNA construct was inserted into the linearized vector pCMV3 (Sino Biological, China). For all cloning steps, the Gibson assembly system was used (Kit NEBuilder HiFi DNA Assembly Master Mix [E2621, NEB, USA]). The final plasmid configuration was: **pCMV3_TMS-fibrillarin_P2A_OFP-myc**.

### Cells transfection and slides preparation

HEp-2 cells were grown to confluence with culture medium DMEM containing 10% fetal bovine serum. The plasmid was transfected with Lipofectamine 3000 Transfection Reagent (L3000008, Invitrogen, USA), diluted in Opti-MEM I Reduced Serum Medium (31985-070, Gibco, USA), following the manufacturer’s protocol. After transfection, cells were seeded in 10-well hydrophobic coated slides (each well with 6-mm diameter) for overnight adherence. After 24h, slides containing the transfected HEp-2 cells were fixed with methanol for 5 min and acetone for 2 min, both cold to -20°C. After air drying, slides were sealed packed and stored at -20°C until use in indirect immunofluorescence assay (IFA).

### CBA IFA reaction

The IFA protocol is similar to a traditional HEp-2 IFA reaction except for the addition of the anti-tag antibody, performed as previously described [41]. Briefly, human sera were diluted 1/80 in PBS containing a mouse anti-Myc monoclonal antibody 9E10 (sc-40, Santa Cruz Biotech), diluted 1/200. Slides containing transfected cells were incubated with this dual antibody mix for 30min at 37°C in a wet chamber and thereafter washed three times for 5min with 0.1% Tween in PBS (PBS-T). Next, two secondary antibodies diluted 1/500 in PBS were incubated simultaneously with the cells at 37°C for 30min in the dark in a wet chamber: anti-human IgG conjugated to Alexa Fluor 488 (A-11013, Invitrogen, USA) and anti-mouse IgG conjugated to Cy3 (715-165-151, Jackson ImmunoResearch, USA). Thereafter slides were washed three times for 5min with PBS-T, assembled with Vectashield containing Dapi (Vector Labs, USA), and covered with coverslips before analysis in a fluorescence microscope (Axio Imager.M2, Carl Zeiss, Germany).

Alternatively for some reactions, a mouse monoclonal anti-fibrillarin antibody was used (clone 72B9) [42] kindly donated by Professor K. Michael Pollard (The Scripps Research Institute, CA, EUA).

### Samples testing

Anti-cell antibody titer and the HEp-2 IFA pattern, including the nucleolar patterns (homogeneous nucleolar AC-8, clumpy nucleolar AC-9 or punctate nucleolar AC-10) were determined using traditional HEp-2 cell IFA slides (Euroimmun, Germany) starting at 1/80 dilution with sequential double dilutions up to end titer.

All samples were submitted to the CBA assay for anti-fibrillarin antibody, as described above, and to other methods for determination of anti-fibrillarin antibodies, as follows. Immunoprecipitation test was performed as previously described [43-45]. Lysate from [^35^S]-methionine-radiolabeled K562 cell (human erythroleukemia) were used for the analysis of the proteins recognized by the serum autoantibodies.

Line blot assay was performed using the Kit Euroline Systemic Sclerosis (Nucleoli) profile (Cat# DL 1532-6401 G, Euroimmun, Germany), following the manufacturer’s protocol. Although this kit can determine the reactivity to other antigens, for this study we only considered the reactivity to the recombinant fibrillarin line blot (Supplementary Figure 5). ELISA analysis was performed using the qualitative Kit for anti-fibrillarin antibody (Cat# MBS701068, MyBioSource, USA), following the manufacturer’s protocol.

### Statistical analysis

Classificatory variables (proportions) were compared with two-tailed Chi squared test. Quantitative and semi-quantitative parameters were tested for Gaussian distribution with “D’Agostino and Pearson normality test”. According to the distribution pattern, they were analyzed by the Mann-Whitney or Student *t*-test. Error bars indicating average and standard deviation (SD) are shown. *P* values were considered significant when below 0.05. All analyses were performed using the software GraphPad Prism 7.0 for Windows.

## Data Availability

All data produced in the present study are available upon reasonable request to the authors.

## References

1. Saketkoo L.A., Frech T., Varju C., Domsic R., Farrell J., et al., A comprehensive framework for navigating patient care in systemic sclerosis: A global response to the need for improving the practice of diagnostic and preventive strategies in SSc. Best Pract Res Clin Rheumatol, 2021. 35(3): p. 101707.

2. Roofeh D. and Khanna D., Management of systemic sclerosis: the first five years. Curr Opin Rheumatol, 2020. 32(3): p. 228–237.

3. Damoiseaux J., Andrade L.E.C., Carballo O.G., Conrad K., Francescantonio P.L.C., et al., Clinical relevance of HEp-2 indirect immunofluorescent patterns: the International Consensus on ANA patterns (ICAP) perspective. Ann Rheum Dis, 2019. 78(7): p. 879–889.

4. Mehra S., Walker J., Patterson K., and Fritzler M.J., Autoantibodies in systemic sclerosis. Autoimmun Rev, 2013. 12(3): p. 340–54.

5. Walker U.A., Tyndall A., Czirjak L., Denton C., Farge-Bancel D., et al., Clinical risk assessment of organ manifestations in systemic sclerosis: a report from the EULAR Scleroderma Trials And Research group database. Ann Rheum Dis, 2007. 66(6): p. 754–63.

6. Shah A.A., Hummers L.K., Casciola-Rosen L., Visvanathan K., Rosen A., et al., Examination of autoantibody status and clinical features associated with cancer risk and cancer-associated scleroderma. Arthritis Rheumatol, 2015. 67(4): p. 1053–61.

7. Kuwana M., Kaburaki J., Okano Y., Tojo T., and Homma M., Clinical and prognostic associations based on serum antinuclear antibodies in Japanese patients with systemic sclerosis. Arthritis Rheum, 1994. 37(1): p. 75–83.

8. Hu P.Q., Fertig N., Medsger T.A., Jr., and Wright T.M., Correlation of serum anti-DNA topoisomerase I antibody levels with disease severity and activity in systemic sclerosis. Arthritis Rheum, 2003. 48(5): p. 1363–73.

9. Steen V.D., Autoantibodies in systemic sclerosis. Semin Arthritis Rheum, 2005. 35(1): p. 35–42.

10. Lazzaroni M.G., Cavazzana I., Colombo E., Dobrota R., Hernandez J., et al., Malignancies in Patients with Anti-RNA Polymerase III Antibodies and Systemic Sclerosis: Analysis of the EULAR Scleroderma Trials and Research Cohort and Possible Recommendations for Screening. J Rheumatol, 2017. 44(5): p. 639–647.

11. Ceribelli A., Cavazzana I., Airo P., and Franceschini F., Anti-RNA polymerase III antibodies as a risk marker for early gastric antral vascular ectasia (GAVE) in systemic sclerosis. J Rheumatol, 2010. 37(7): p. 1544.

12. Hamaguchi Y., Kodera M., Matsushita T., Hasegawa M., Inaba Y., et al., Clinical and immunologic predictors of scleroderma renal crisis in Japanese systemic sclerosis patients with anti-RNA polymerase III autoantibodies. Arthritis Rheumatol, 2015. 67(4): p. 1045–52.

13. Patterson K.A., Roberts-Thomson P.J., Lester S., Tan J.A., Hakendorf P., et al., Interpretation of an Extended Autoantibody Profile in a Well-Characterized Australian Systemic Sclerosis (Scleroderma) Cohort Using Principal Components Analysis. Arthritis Rheumatol, 2015. 67(12): p. 3234–44.

14. Aggarwal R., Lucas M., Fertig N., Oddis C.V., and Medsger T.A., Jr., Anti-U3 RNP autoantibodies in systemic sclerosis. Arthritis Rheum, 2009. 60(4): p. 1112–8.

15. Sharif R., Fritzler M.J., Mayes M.D., Gonzalez E.B., McNearney T.A., et al., Anti-fibrillarin antibody in African American patients with systemic sclerosis: immunogenetics, clinical features, and survival analysis. J Rheumatol, 2011. 38(8): p. 1622–30.

16. Kayser C. and Fritzler M.J., Autoantibodies in systemic sclerosis: unanswered questions. Front Immunol, 2015. 6: p. 167.

17. Chan E.K.L., von Muhlen C.A., Fritzler M.J., Damoiseaux J., Infantino M., et al., The International Consensus on ANA Patterns (ICAP) in 2021-The 6th Workshop and Current Perspectives. J Appl Lab Med, 2022. 7(1): p. 322–330.

18. Reveille J.D., Fischbach M., McNearney T., Friedman A.W., Aguilar M.B., et al., Systemic sclerosis in 3 US ethnic groups: a comparison of clinical, sociodemographic, serologic, and immunogenetic determinants. Semin Arthritis Rheum, 2001. 30(5): p. 332–46.

19. Ferri C., Bernini L., Cecchetti R., Latorraca A., Marotta G., et al., Cutaneous and serologic subsets of systemic sclerosis. J Rheumatol, 1991. 18(12): p. 1826–32.

20. Bunn C.C., Denton C.P., Shi-Wen X., Knight C., and Black C.M., Anti-RNA polymerases and other autoantibody specificities in systemic sclerosis. Br J Rheumatol, 1998. 37(1): p. 15–20.

21. Hesselstrand R., Scheja A., Shen G.Q., Wiik A., and Akesson A., The association of antinuclear antibodies with organ involvement and survival in systemic sclerosis. Rheumatology (Oxford), 2003. 42(4): p. 534–40.

22. Nandiwada S.L., Peterson L.K., Mayes M.D., Jaskowski T.D., Malmberg E., et al., Ethnic Differences in Autoantibody Diversity and Hierarchy: More Clues from a US Cohort of Patients with Systemic Sclerosis. J Rheumatol, 2016. 43(10): p. 1816–1824.

23. Satoh M., Ceribelli A., Hasegawa T., and Tanaka S., Clinical Significance of Antinucleolar Antibodies: Biomarkers for Autoimmune Diseases, Malignancies, and others. Clin Rev Allergy Immunol, 2022.

24. Stochmal A., Czuwara J., Trojanowska M., and Rudnicka L., Antinuclear Antibodies in Systemic Sclerosis: an Update. Clin Rev Allergy Immunol, 2020. 58(1): p. 40–51.

25. Mahler M., Kim G., Roup F., Bentow C., Fabien N., et al., Evaluation of a novel particle-based multi-analyte technology for the detection of anti-fibrillarin antibodies. Immunol Res, 2021. 69(3): p. 239–248.

26. Peterson L.K., Jaskowski T.D., Mayes M.D., and Tebo A.E., Detection of anti-U3-RNP/fibrillarin IgG antibodies by line immunoblot assay has comparable clinical significance to immunoprecipitation testing in systemic sclerosis. Immunol Res, 2016. 64(2): p. 483–8.

27. van den Hoogen F., Khanna D., Fransen J., Johnson S.R., Baron M., et al., 2013 classification criteria for systemic sclerosis: an American college of rheumatology/European league against rheumatism collaborative initiative. Ann Rheum Dis, 2013. 72(11): p. 1747–55.

28. Molina R.D., Conzatti L.P., da Silva A.P.B., Goi L.D.S., da Costa B.K., et al., Detection of autoantibodies in central nervous system inflammatory disorders: Clinical application of cell-based assays. Mult Scler Relat Disord, 2020. 38: p. 101858.

29. Hoxha E., Beck L.H., Jr., Wiech T., Tomas N.M., Probst C., et al., An Indirect Immunofluorescence Method Facilitates Detection of Thrombospondin Type 1 Domain-Containing 7A-Specific Antibodies in Membranous Nephropathy. J Am Soc Nephrol, 2017. 28(2): p. 520–531.

30. Hoxha E., Harendza S., Zahner G., Panzer U., Steinmetz O., et al., An immunofluorescence test for phospholipase-A(2)-receptor antibodies and its clinical usefulness in patients with membranous glomerulonephritis. Nephrol Dial Transplant, 2011. 26(8): p. 2526–32.

31. Satoh M., Chan E.K., Ho L.A., Rose K.M., Parks C.G., et al., Prevalence and sociodemographic correlates of antinuclear antibodies in the United States. Arthritis Rheum, 2012. 64(7): p. 2319–27.

32. Dinse G.E., Parks C.G., Weinberg C.R., Co C.A., Wilkerson J., et al., Increasing Prevalence of Antinuclear Antibodies in the United States. Arthritis Rheumatol, 2020. 72(6): p. 1026–1035.

33. Sener A.G., Afsar I., and Demirci M., Evaluation of antinuclear antibodies by indirect immunofluorescence and line immunoassay methods’: four years’ data from Turkey. APMIS, 2014. 122(12): p. 1167–70.

34. Mengeloglu Z., Tas T., Kocoglu E., Aktas G., and Karabork S., Determination of Anti-nuclear Antibody Pattern Distribution and Clinical Relationship. Pak J Med Sci, 2014. 30(2): p. 380–3.

35. Arnett F.C., Reveille J.D., Goldstein R., Pollard K.M., Leaird K., et al., Autoantibodies to fibrillarin in systemic sclerosis (scleroderma). An immunogenetic, serologic, and clinical analysis. Arthritis Rheum, 1996. 39(7): p. 1151–60.

36. Tormey V.J., Bunn C.C., Denton C.P., and Black C.M., Anti-fibrillarin antibodies in systemic sclerosis. Rheumatology (Oxford), 2001. 40(10): p. 1157–62.

37. Kuwana M., Circulating Anti-Nuclear Antibodies in Systemic Sclerosis: Utility in Diagnosis and Disease Subsetting. J Nippon Med Sch, 2017. 84(2): p. 56–63.

38. de Oliveira S.M., Martins L.V.O., Lupino-Assad A.P., Medeiros-Ribeiro A.C., de Moraes D.A., et al., Severity and mortality of COVID-19 in patients with systemic sclerosis: a Brazilian multicenter study. Semin Arthritis Rheum, 2022. 55: p. 151987.

39. Galie N., Humbert M., Vachiery J.L., Gibbs S., Lang I., et al., 2015 ESC/ERS Guidelines for the diagnosis and treatment of pulmonary hypertension: The Joint Task Force for the Diagnosis and Treatment of Pulmonary Hypertension of the European Society of Cardiology (ESC) and the European Respiratory Society (ERS): Endorsed by: Association for European Paediatric and Congenital Cardiology (AEPC), International Society for Heart and Lung Transplantation (ISHLT). Eur Respir J, 2015. 46(4): p. 903–75.

40. Lau E.M., Tamura Y., McGoon M.D., and Sitbon O., The 2015 ESC/ERS Guidelines for the diagnosis and treatment of pulmonary hypertension: a practical chronicle of progress. Eur Respir J, 2015. 46(4): p. 879–82.

41. Keppeke G.D., Prado M.S., Nunes E., Perazzio S.F., Rodrigues S.H., et al., Differential capacity of therapeutic drugs to induce Rods/Rings structures in vitro and in vivo and generation of anti-Rods/Rings autoantibodies. Clin Immunol, 2016. 173: p. 149–156.

42. Reimer G., Pollard K.M., Penning C.A., Ochs R.L., Lischwe M.A., et al., Monoclonal autoantibody from a (New Zealand black x New Zealand white)F1 mouse and some human scleroderma sera target an Mr 34,000 nucleolar protein of the U3 RNP particle. Arthritis Rheum, 1987. 30(7): p. 793–800.

43. Keppeke G.D., Satoh M., Ferraz M.L., Chan E.K., and Andrade L.E., Temporal evolution of human autoantibody response to cytoplasmic rods and rings structure during anti-HCV therapy with ribavirin and interferon-alpha. Immunol Res, 2014. 60(1): p. 38–49.

44. Carcamo W.C., Ceribelli A., Calise S.J., Krueger C., Liu C., et al., Differential reactivity to IMPDH2 by anti-rods/rings autoantibodies and unresponsiveness to pegylated interferon-alpha/ribavirin therapy in US and Italian HCV patients. J Clin Immunol, 2013. 33(2): p. 420–6.

45. Satoh M., Langdon J.J., Hamilton K.J., Richards H.B., Panka D., et al., Distinctive immune response patterns of human and murine autoimmune sera to U1 small nuclear ribonucleoprotein C protein. J Clin Invest, 1996. 97(11): p. 2619–26.

