## Supplementary material for "A Cell-Based Assay for Detection of Anti-Fibrillarin Autoantibodies in Systemic Sclerosis": Suppl. Figure

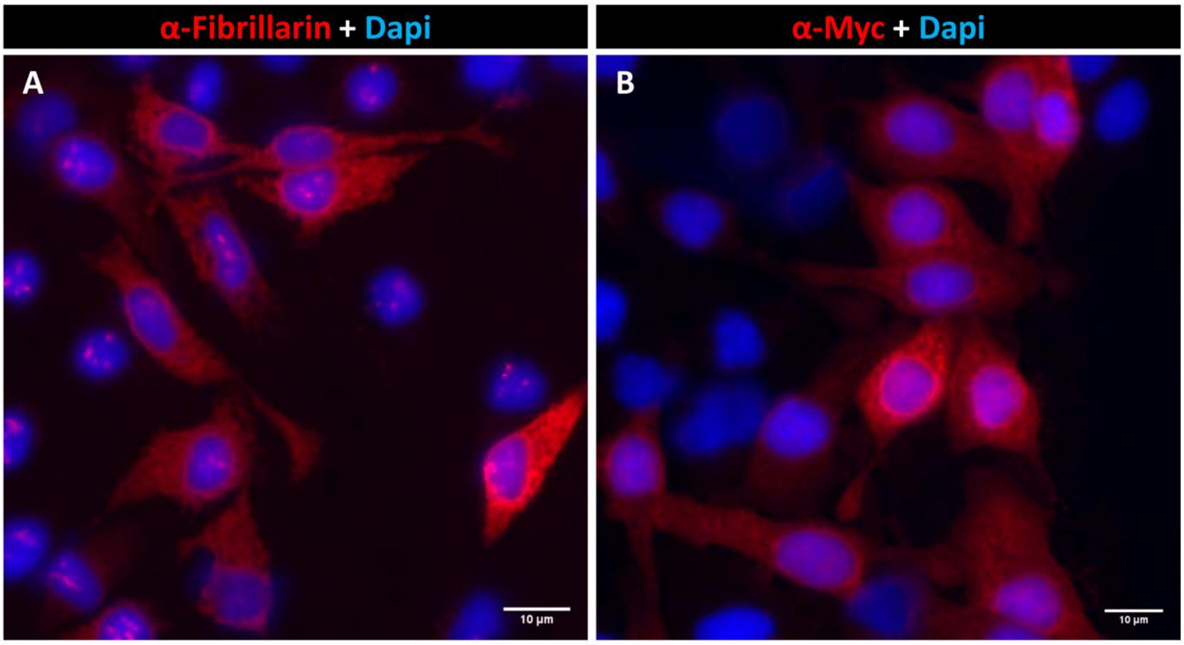


**Suppl. Figure 1. TMS-Fibrillarin was relocated to the cell membrane.** HEp-2 cells transfected with TMS-Fibrillarin_P2A_OFP-myc were probed with mouse monoclonal antibodies. (A) Fibrillarin labeling in the cytoplasmic membrane. (B) Anti-Myc antibody label cells expressing TMS-Fibrillarin. Scale bar = 10µm.

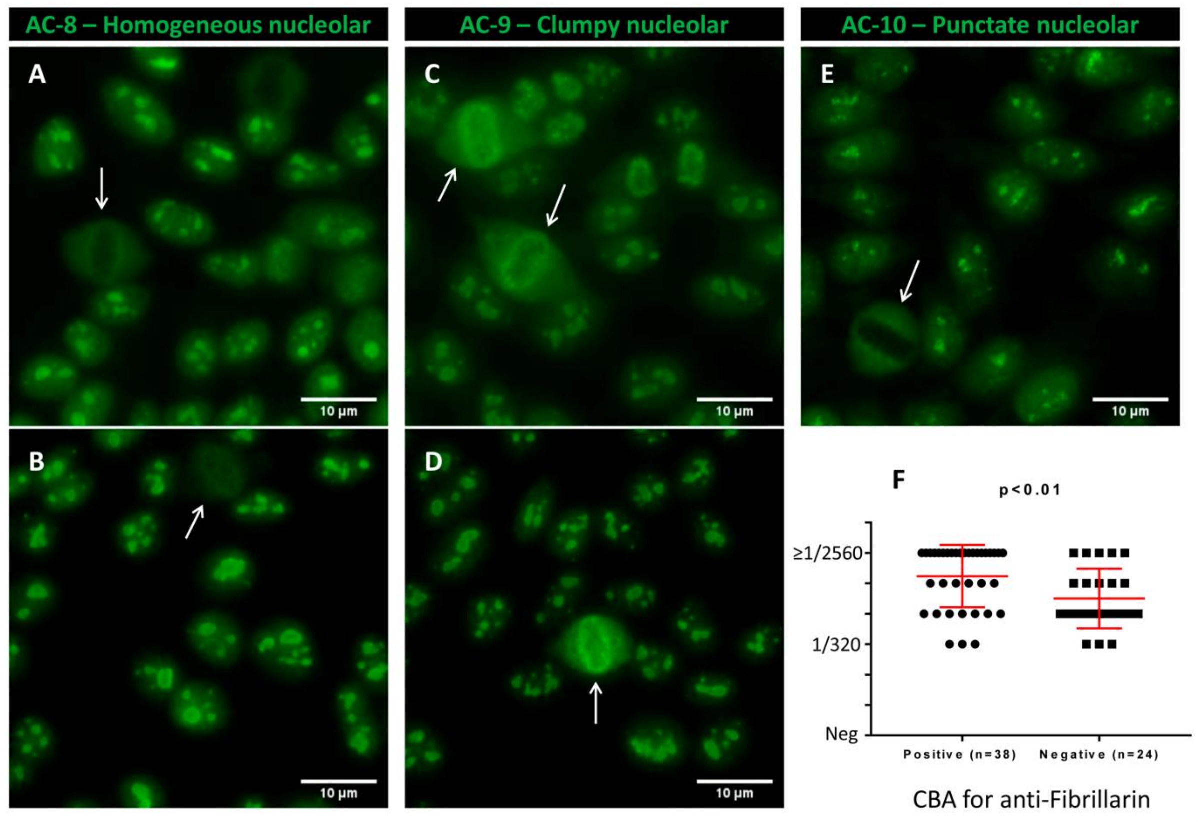


**Suppl. Figure 2. Different nucleolar patterns in the HEp-2 IFA test.** Example of samples presenting Homogenous nucleolar AC-8 (A – B), Clumpy nucleolar AC-9 (C – D) and Punctate nucleolar AC-10 (E). The peri-chromosomal staining at the metaphase plates were considered for classification as Clumpy nucleolar. Arrows in all panels indicate metaphase plates. Scale bar = 10µm. (F) Average HEp-2 IFA titer in samples positive or negative for anti-fibrillarin. Error bars indicate mean ± S.D.

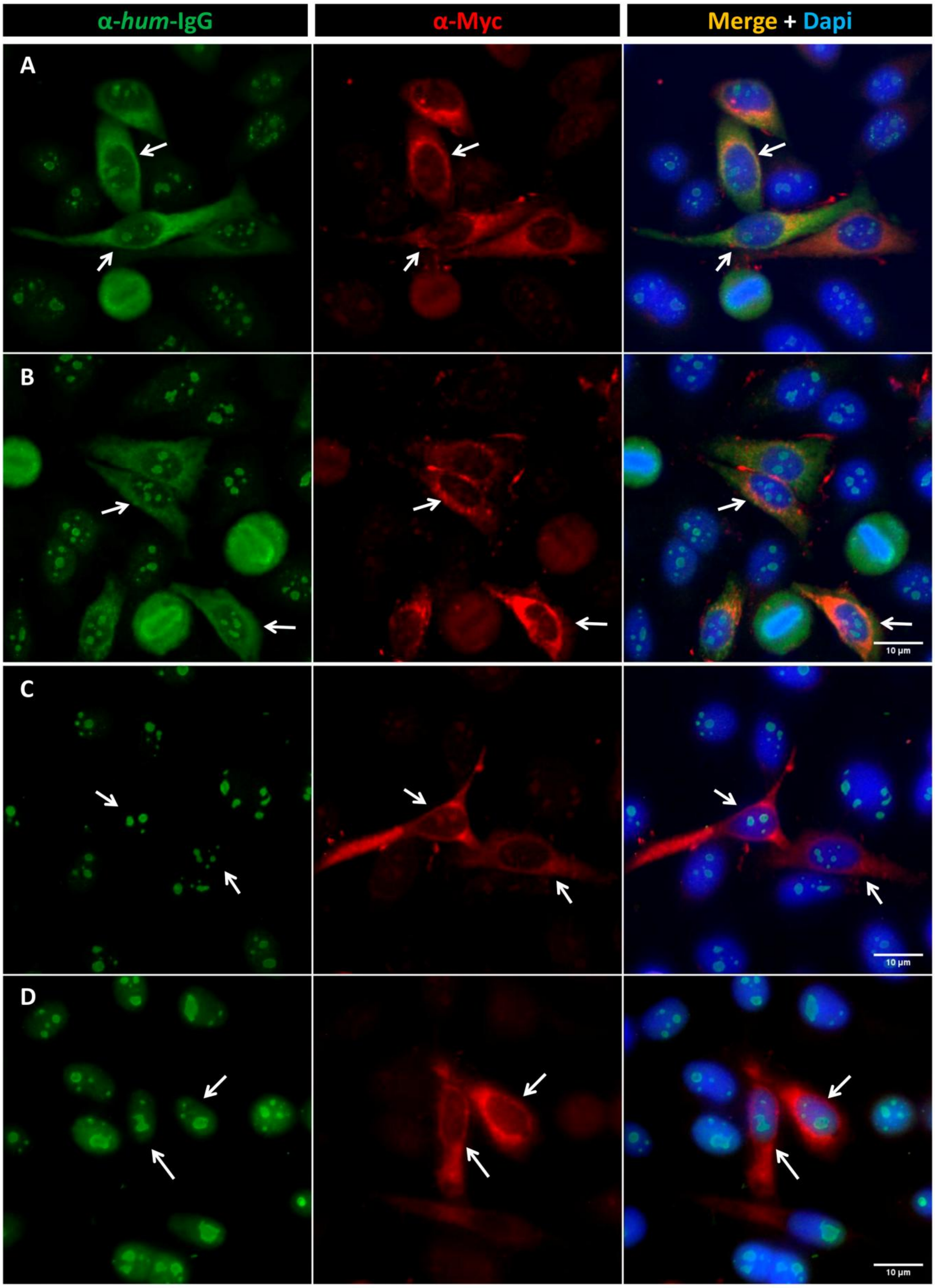


**Suppl. Figure 3. Examples of positive and negative results in the Fibrillarin/CBA.** (A-B) Examples of samples with positive labeling of TMS-Fibrillarin. (C-D) Examples of samples with negative labeling of TMS-Fibrillarin. Arrows indicate cells expressing the TMS-Fibrillarin. Scale bar = 10µm.

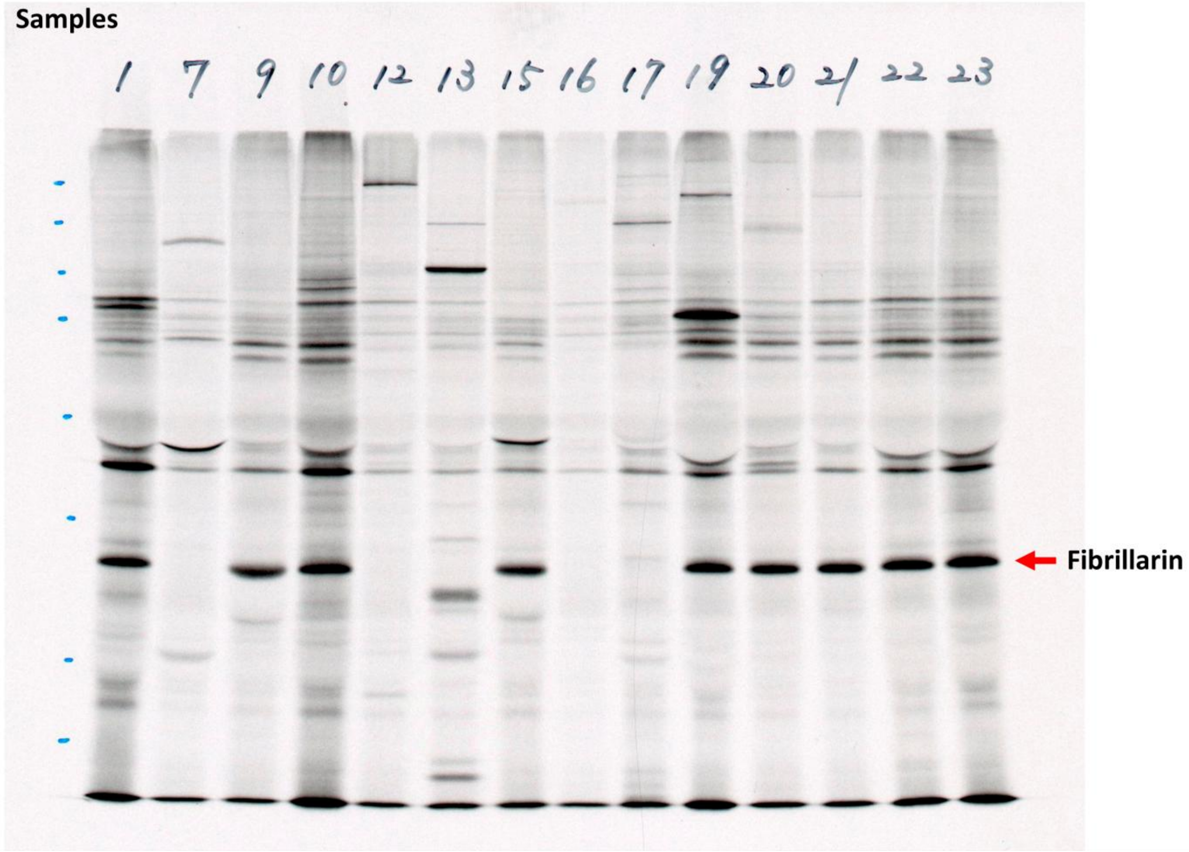


**Suppl. Figure 4. Example of immunoprecipitation.** Example (1 gel = 14 samples) indicating the ~34kDa fibrillarin band.

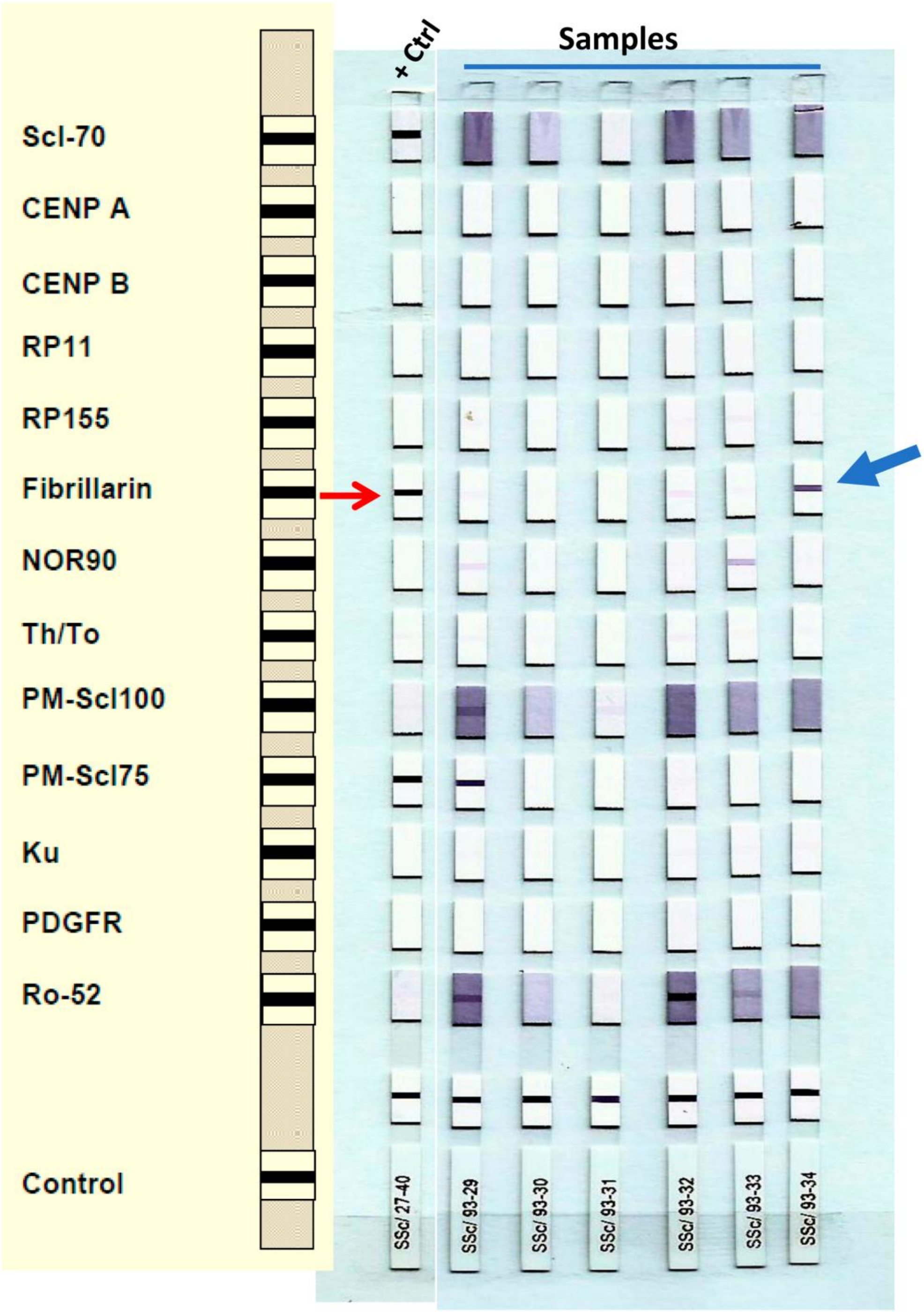


**Suppl Figure 5. Example of line blot assay.** For the line blot analyses, Euroline Systemic sclerosis (Nucleoli) profile was used. Red arrow indicates the fibrillarin line. Blue arrow indicates a positive sample.
